# The international and intercontinental spread and expansion of antimicrobial-resistant *Salmonella* Typhi

**DOI:** 10.1101/2021.09.03.21262852

**Authors:** Kesia Esther da Silva, Arif Mohammad Tanmoy, Agila Kumari Pragasam, Junaid Iqbal, Mohammad Saiful Islam Sajib, Ankur Mutreja, Balaji Veeraraghavan, Dipesh Tamrakar, Farah Naz Qamar, Gordon Dougan, Isaac Bogoch, Jessica C Seidman, Jivan Shakya, Krista Vaidya, Megan E. Carey, Rajeev Shrestha, Seema Irfan, Stephen Baker, Steve P. Luby, Yanjia Cao, Zoe Anne Dyson, Denise O. Garrett, Jacob John, Gagandeep Kang, Yogesh Hooda, Samir K. Saha, Senjuti Saha, Jason R. Andrews

## Abstract

The emergence of increasingly antimicrobial-resistant (AMR) *Salmonella enterica* serovar Typhi (*S*. Typhi) threatens to undermine effective treatment and control. Here, aiming to investigate the temporal and geographic patterns of emergence and spread of AMR *S*. Typhi, we sequenced 3,489 *S*. Typhi isolated from prospective surveillance in South Asia and combined these with a global collection of 4,169 *S*. Typhi genomes. Our analysis revealed that independent acquisition of plasmids and homoplastic mutations conferring AMR have occurred repeatedly in multiple lineages of *S*. Typhi, predominantly arising in South Asia. We found evidence of frequent international and intercontinental transfers of AMR *S*. Typhi, followed by rapid expansion and replacement of antimicrobial-susceptible clades.

## Introduction

Typhoid fever, the disease caused by *Salmonella enterica* serovar Typhi (*S*. Typhi), remains a major worldwide public health concern (1). The organism causes 11 million illnesses and >100,000 deaths annually (2,3). The highest typhoid fever incidence rates occur in South Asia, which contains 70% of the global disease burden, but substantial morbidity and mortality also occur in sub-Saharan Africa, Southeast Asia, and Oceania. In the pre-antimicrobial era, the case fatality rate of typhoid fever was around 15%, but morbidity and mortality declined dramatically following the introduction of chloramphenicol in the 1940s, followed by ampicillin and trimethoprim-sulfamethoxazole in the 1960s (4,5).

The effectiveness of antimicrobial therapy for typhoid fever has been repeatedly threatened by the emergence and expansion of organisms that exhibit resistance to the principal antimicrobials. Multi-drug resistant (MDR) variants, harboring genes encoding resistance to ampicillin, chloramphenicol, and trimethoprim-sulfamethoxazole first emerged in the 1970s; subsequently, a single lineage (H58; 4.3.1) was introduced into sub-Saharan Africa and Southeast Asia from South Asia and became globally dominant (6,7). The genes facilitating resistance to these specific antimicrobials were originally only located on IncHI1 plasmids but later also became integrated into the chromosome. The fluoroquinolones were initially highly effective against early MDR *S*. Typhi organisms and became the mainstay of therapy in South Asia in the 1990s. However, fluoroquinolone non-susceptible isolates started to emerge in the mid 1990s and, within 20 years, the majority of *S*. Typhi in South Asia contained mutations in the quinolone resistance-determining regions (QRDR) (4) (8). In 2016, a large outbreak of *S*. Typhi containing plasmid-mediated resistance to third generation cephalosporins and fluoroquinolones and chromosomally located genes encoding the MDR phenotype was identified in Pakistan and termed extensively drug-resistant (XDR) (9). More recently, a single polymorphism in the AcrB efflux pump conferring resistance to azithromycin was found to have independently arisen in multiple lineages of *S*. Typhi, threatening the efficacy of all oral antibiotic antimicrobials for typhoid treatment (10). Taken together, *S*. Typhi has exhibited resistance to all oral drugs known to be effective for its treatment, although it has not yet been identified in the same clone.

Typhoid conjugate vaccines (TCV) have recently proven effective for disease prevention, and the World Health Organization recommends prioritized introduction in countries with a high burden of antimicrobial resistant *S*. Typhi (11). However, given the current trajectory of antimicrobial resistant (AMR) in *S*. Typhi waiting until a high AMR burden is present within a country to introduce typhoid vaccines may ill-advised. Understanding the historical emergence, expansion and geographic spread of antimicrobial resistant *S*. Typhi may yield insights into where resistant strains might spread and how quickly they will become dominant within populations.

Here, we leveraged prospective, population-based typhoid surveillance studies from four of the highest burden countries in South Asia: Bangladesh, India, Nepal and Pakistan. We sequenced 3,489 *S*. Typhi organisms isolated over a six-year period, and these data were combined with a global collection of >4,000 additional genomes sequences to investigate the emergence and geographical spread of AMR *S*. Typhi over the past three decades.

## Results

### Genotypic diversity and phylogenetic structure of S. Typhi in South Asia

A total of 3,489 *S*. Typhi isolates from four countries (Bangladesh, India, Nepal, and Pakistan) collected between 2014 and 2019 were sequenced. Genotype analysis identified 29 distinct genotypes (**Figure S1**). The majority of isolates (2,474; 70.9%) belonged to genotype 4.3.1 (haplotype H58). The major sublineages of H58 (lineage I and lineage II) were common, with H58 lineage I (genotype 4.3.1.1) dominant in Bangladesh and Pakistan, and H58 lineage II (genotype 4.3.1.2) the most common genotype in India and Nepal. Among non-H58 isolates, the most common subclades were subclade 3.2.2 (190; 5.5%), 3.3.2 (161; 4.6%), 2.3.3 (140; 4.0%), 2.5 (123; 3.5%), and 3.3.1 (85; 2.4%).

We identified multiple, phylogenetically linked sub-lineages shared across South Asia (**Figure S2**), most regularly between Bangladesh, Nepal, and India. Within the H58 isolates, *S*. Typhi 4.3.1.2 isolates formed distinct clades with intermingled isolates from India and Nepal, while 4.3.1.3 isolates, identified predominantly in Bangladesh, clustered with few isolates from India. In contrast, the H58 isolates from Pakistan largely clustered independently and was dominated by a monophyletic XDR clade (4.3.1.1.P1).

### Global phylogenetic structure and determinants of antimicrobial resistance

To provide additional context for the 3,489 *S*. Typhi new South Asian genome sequences, and better understand temporal and spatial distribution of lineages and antimicrobial resistance, we constructed a further phylogeny incorporating an additional 4,169 *S*. Typhi sequences from organisms isolated from 1905 to 2018 from >70 countries (**Figure 1**). Overall, the new sequences clustered with previously sequenced South Asian isolates, generating distinct geographic structure. Genotype 4.3.1 formed a large subclade. Primary clades 2, 3 and 4 were distributed across continents with limited isolates outside these clades. Notably, four subclades (2.3.3, 2.5, 3.2.2, and 3.3) were dominant in South Asia, accounting for 75.7% of all non-H58 organisms.

**Figure 1.**
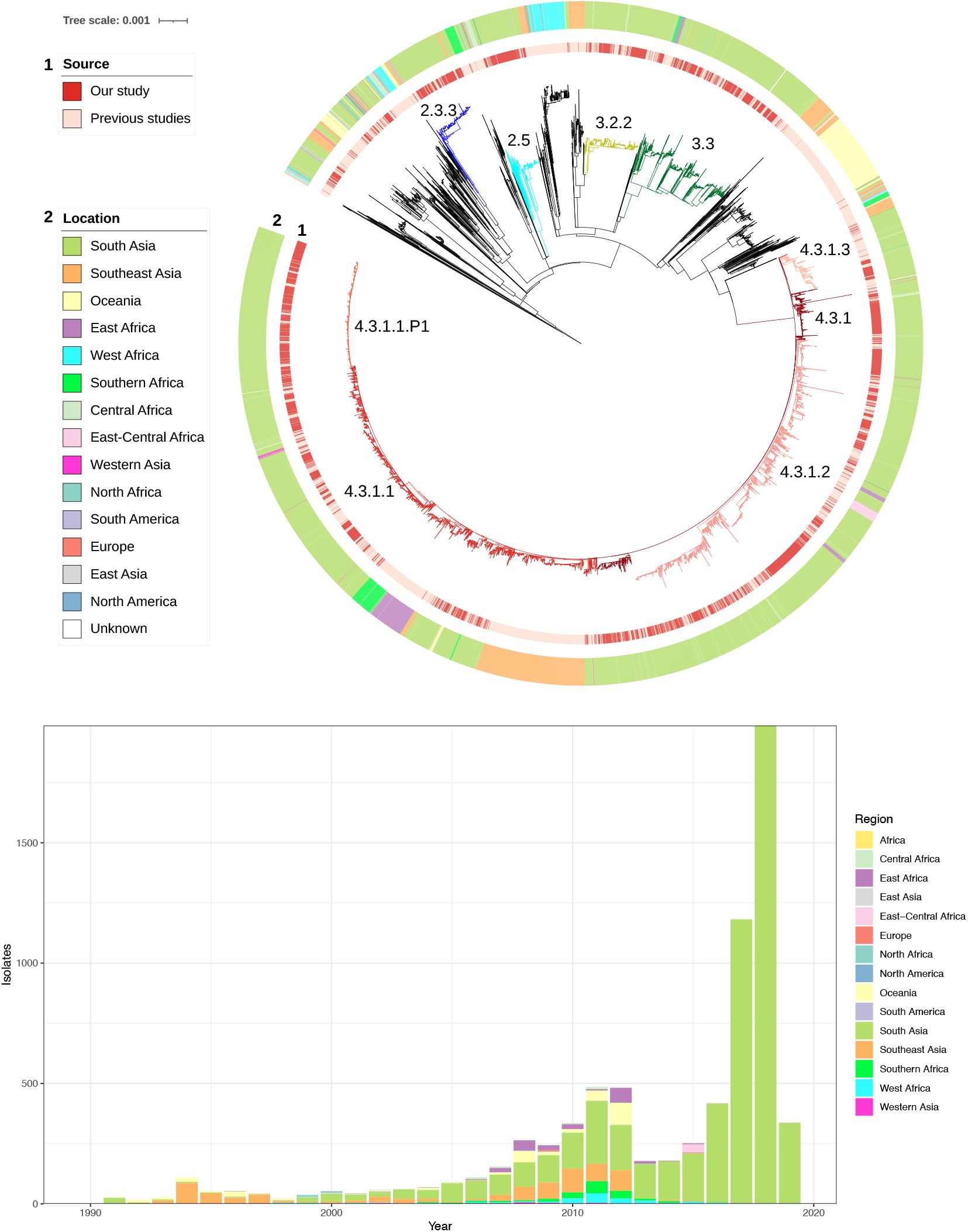
Global phylogeny of *Salmonella* Typhi. **(a)** Maximum likelihood tree of 7,658 *S*. Typhi isolates from the global collection. Branch colors indicate the lineages 2.3.3 (blue), 2.5 (turquoise), 3.2.2 (yellow), 3.3 (green), 4.3.1 (dark red), 4.3.1.1 (red), 4.3.1.1.P1 (orange), 4.3.1.2 (pink), 4.3.1.3 (salmon) and other non-H58 (black). The inner ring indicates the source. The outer ring indicates the region of isolation. The scale bar indicates nucleotide substitutions per site **(b)** Temporal distribution of sequenced *S*. Typhi isolates by region.

We classified isolates as multidrug-resistant if they containing genes conferring resistance to ampicillin (*bla*_TEM-1_), chloramphenicol (*catA1*), and trimethoprim-sulfamethoxazole (*dfrA7, sul1*, or *sul2*). From 2000 onwards, we observed a declining trend in MDR isolates in Bangladesh and India, a stable low proportion (<5%) in Nepal, and an increasing proportion in Pakistan and Africa (**Figure S3a**). Acquired resistance genes that contribute to the MDR phenotype were identified in 26.8% of the isolates of the global genome collection, including 98.4% of the H58 isolates, in comparison to just 1.6% of the non-H58 isolates (1.1.1, 2.5.1, 3.1.1, 3.2.1, 3.4, 4). Among these rare non-H58 multidrug-resistant isolates, resistance was almost exclusively entirely plasmid-mediated (96.9%). In contrast, for H58 isolates we observed that plasmid-mediated resistance was persistent in the H58 isolates in the 1990s, but from 2000 onward was less frequent, with most MDR isolates containing chromosomal insertions of drug-resistance genes (75.2%).

In contrast to the temporal trends in MDR, there was a consistent rise in the proportion of global *S*. Typhi that were fluoroquinolone non-susceptible (FQNS), primarily associated with mutations *gryA, gyrB, parC*, and *parE*) (**Figure S3b**). The most dramatic increase in FQNS *S*. Typhi occurred in Bangladesh, exceeding 85% by early 2000, followed by India, Pakistan, and Nepal, reaching >95% in all four South Asian countries. FQNS *S*. Typhi increased from <20% in 2007 year to >60% by 2011 in Southeast Asia. In Africa, this increase occurred more recently, originating in 2010. Overall, we found that QRDR mutations were significantly more common in the H58 isolates (89.1%) compared with other lineages (45.4%) (*p*<0.0001). From 2010 onwards, an increasing number of isolates containing multiple QRDR mutations; >10% of all isolates harbored three mutations (**Figure S3**). Among the novel genome sequences, 437 were ‘triple mutants’ (Table S3), which are highly associated with full resistance and failure to respond to fluoroquinolone therapy (12). The majority (402/437; 92%) of these organisms occurred in H58 lineage II (4.3.1.2) in India and Nepal; the second most common (15/437; 3.4%) triple mutant genotype was 3.3 and predominantly isolated in India.

Susceptibility to fluoroquinolones can be further reduced via plasmid-mediated acquisition of *qnr* genes. We identified *qnrS* in two non-H58 isolates (genotype 3) and 686 H58 isolates that included genotype 4.3.1 (n = 3), 4.3.1.1 (n = 5), 4.3.1.P1 (n = 550), and 4.3.1.3 (n = 125). Most H58 lineage I isolates from Pakistan were XDR (4.3.1.P1) carrying the previously identified composite transposon containing *bla*_TEM-1_, *catA1, dfrA7, sul1, sul2* inserted in the chromosome, and *bla*_CTX-M-15_ and *qnrS* associated with an IncY plasmid (9). Azithromycin resistance, conferred by *acrB* mutations (R717Q and R717L), was identified in 54 isolates across eight different genotypes including genotype 4.3.1 (n = 1), 4.3.1.1 (n = 31), 4.3.1.2 (n = 5), 4.3.1.3 (n = 2), and non-H58 isolates comprising, genotype 2.3.3 (n = 2), 3.2.2 (n = 9), 3.3.2 (n = 3) and 3.5.4 (n = 1).

### Antimicrobial resistance and growth of the effective population size

To investigate how the AMR has shaped the effective population size of *S*. Typhi, we generated timed phylogenies and modeled the effective population size of antimicrobial susceptible and antimicrobial resistant organisms over time. To minimize the effect of location and lineage, we focused on the largest haplotype (H58) and performed analyses within countries, evaluating key AMR determinants. In Nepal, we found that the effective population size (N_e_) of *S*. Typhi containing one or two QRDR mutations rose steadily from 2000, beginning to decline from 2017, while triple mutants have steadily increased from 2010 (**Figure 2)**. In Pakistan, the N_e_ of non-XDR H58 *S*. Typhi increased from 2000 until around 2015 and began to fall; XDR organisms emerged and have been rapidly growing in frequency since 2016, eclipsing the effective population of non-XDR organisms by 2018. In Bangladesh, the N_e_ of H58 had slowly declined from around 2010; however, azithromycin-resistant organisms emerged in 2013 with a corresponding increase in N_e_. In all three settings, organisms with key AMR conferring mutations or genes appear to be replacing their susceptible (or, in the case of fluoroquinolones, less-resistant) counterparts.

**Figure 2.**
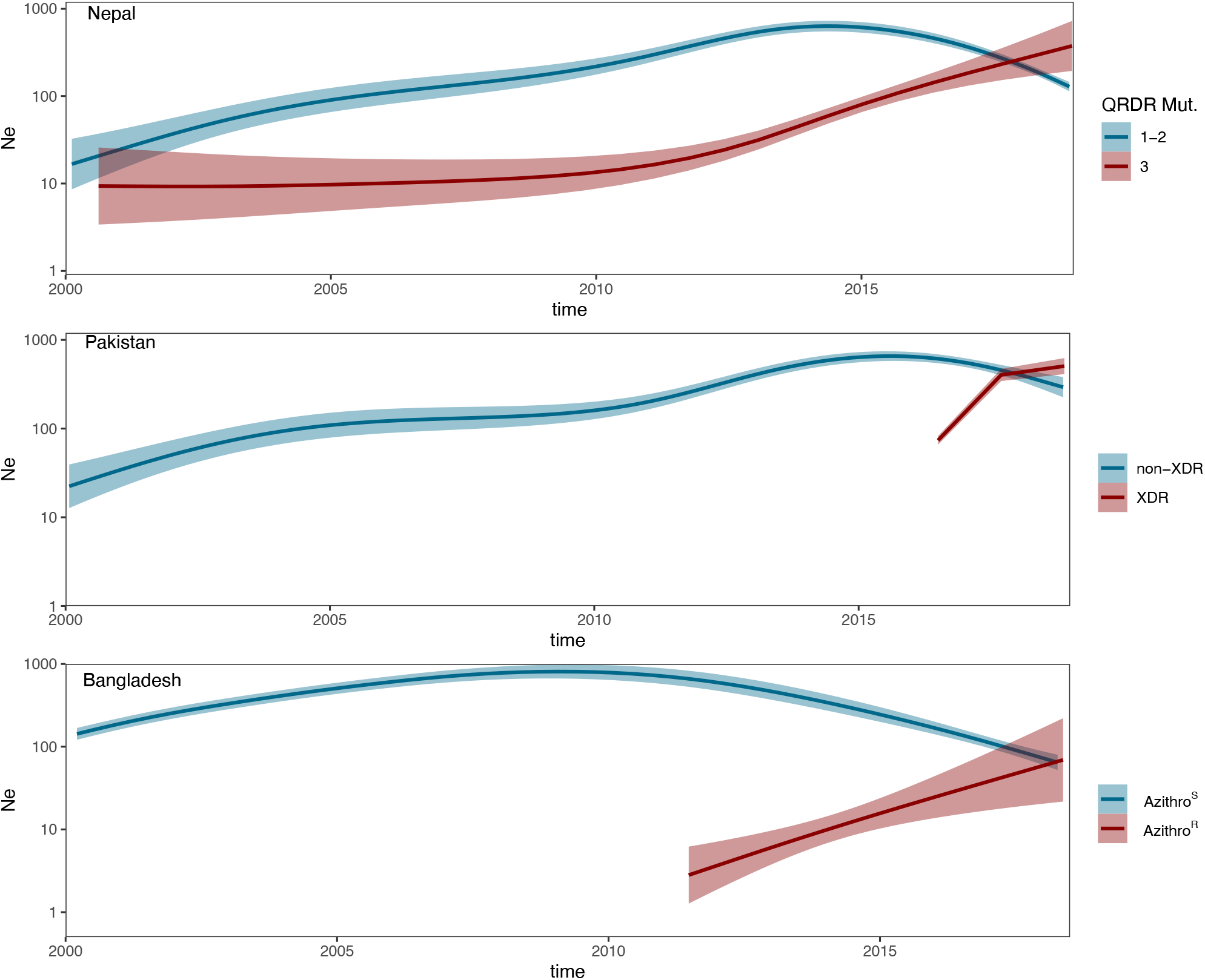
The effective population size (N_e_) of H58 lineages according to antimicrobial resistance genotype in Nepal, Pakistan and Bangladesh. In Nepal, strains containing 1-2 mutations in the quinolone resistance determining region (QRDR) were compared with those containing 3 mutations. In Pakistan, extensively drug-resistant (XDR) strains were compared with non-XDR strains. In Bangladesh, strains containing *acrB* mutations conferring azithromycin-resistance were compared with those not containing the mutations. Light shading represents the 95% high probability density (HPD) intervals of the estimates.

### The global phylogeography of S. Typhi

Using country of sampling as a discrete trait, we generated dated phylogenies to reconstruct the evolutionary history and geographic spread of H58 lineage and the four 4 common non-H58 genotypes. These five genotypes accounted for 75% of all isolates from the past decade. The mean nucleotide substitution rates for each lineage are described in **Table S4**.

Phylogeographic reconstruction of H58 isolates (4.3.1, 4.3.1.1, 4.3.1.1.P1, 4.3.2.1 and 4.3.1.3) estimated that the time of most recent common ancestor (tMRCA) of all contemporary H58 *S*. Typhi strains existed around 37 years ago (1984). The distribution of isolates and the tree topology are consistent with at least 138 international transfer events, including multiple introductions within South Asia and dissemination from South Asia into Southeast Asia and Africa, as well as many travel-related cases identified in the U.K. and U.S. (**Figure 3, Figure 4**). The distribution of QRDR mutations within the phylogeny demonstrated that these resistance-conferring mutations have arisen independently on at least 80 distinct occasions. We also predicted that ciprofloxacin-resistant triple mutant isolates most likely originated in India around 1996 and were introduced into Pakistan between 2005 and 2013 and into Nepal on at least three occasions (2003-2015). Furthermore, we identified frequent transmissions of international transfer of MDR isolates (n = 33), with multiple introductions from South Asia followed by local expansion. In addition, the phylogeographic analyses also showed that the Pakistani XDR lineage (genotype 4.3.1.1.P1) emerged around 2015 and has been subsequently identified on multiple occasions in United Kingdom and United States.

**Figure 3.**
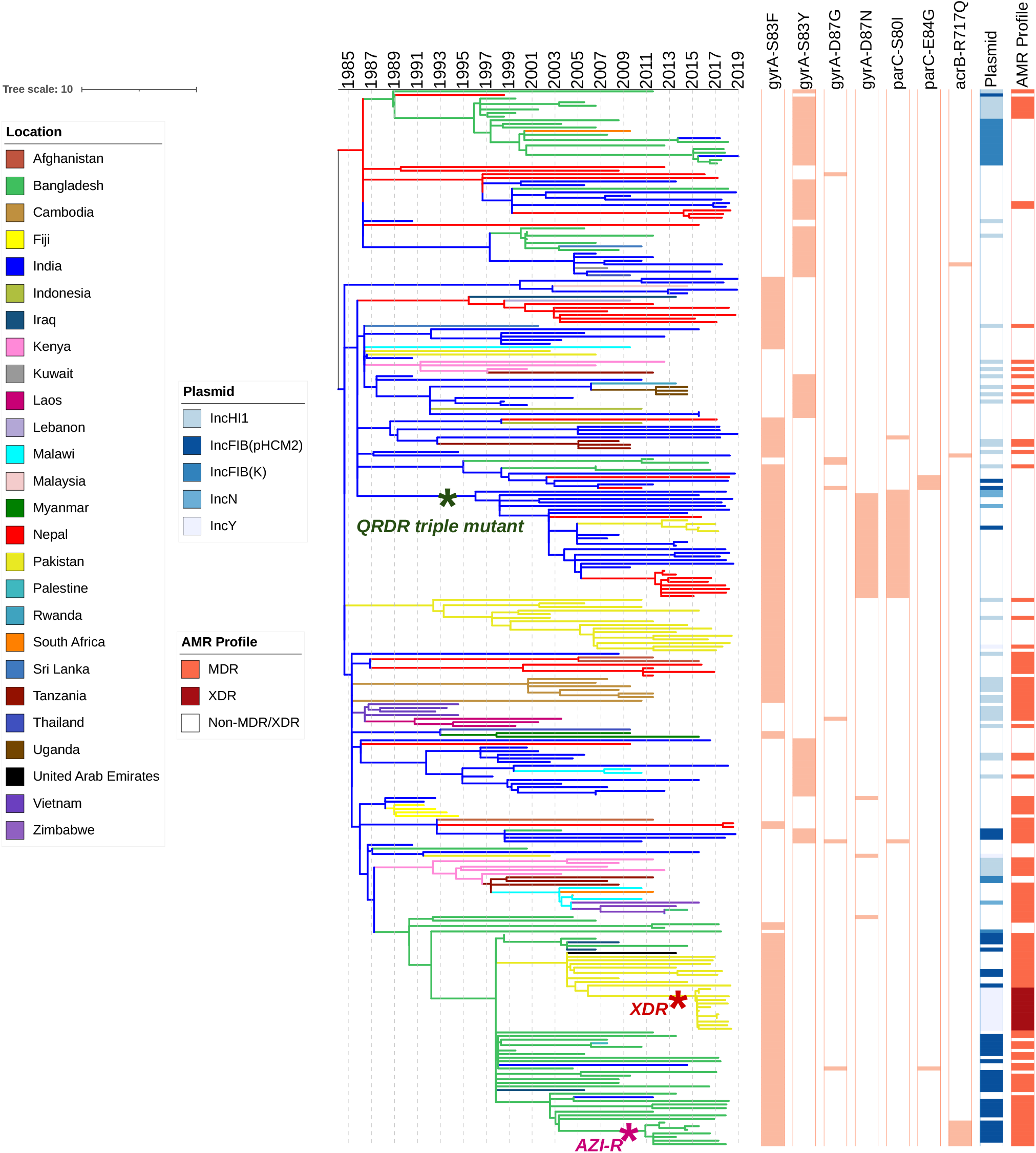
Phylogeography and global expansion of genotype 4.3.1 (H58) *S*. Typhi isolates. Timed phylogenetic tree of genotype 4.3.1 *S*. Typhi isolates. The branch lengths are scaled in years and are colored according to the location of the most probable ancestor of descendant nodes. The scale bar indicates nucleotide substitutions per site.

**Figure 4.**
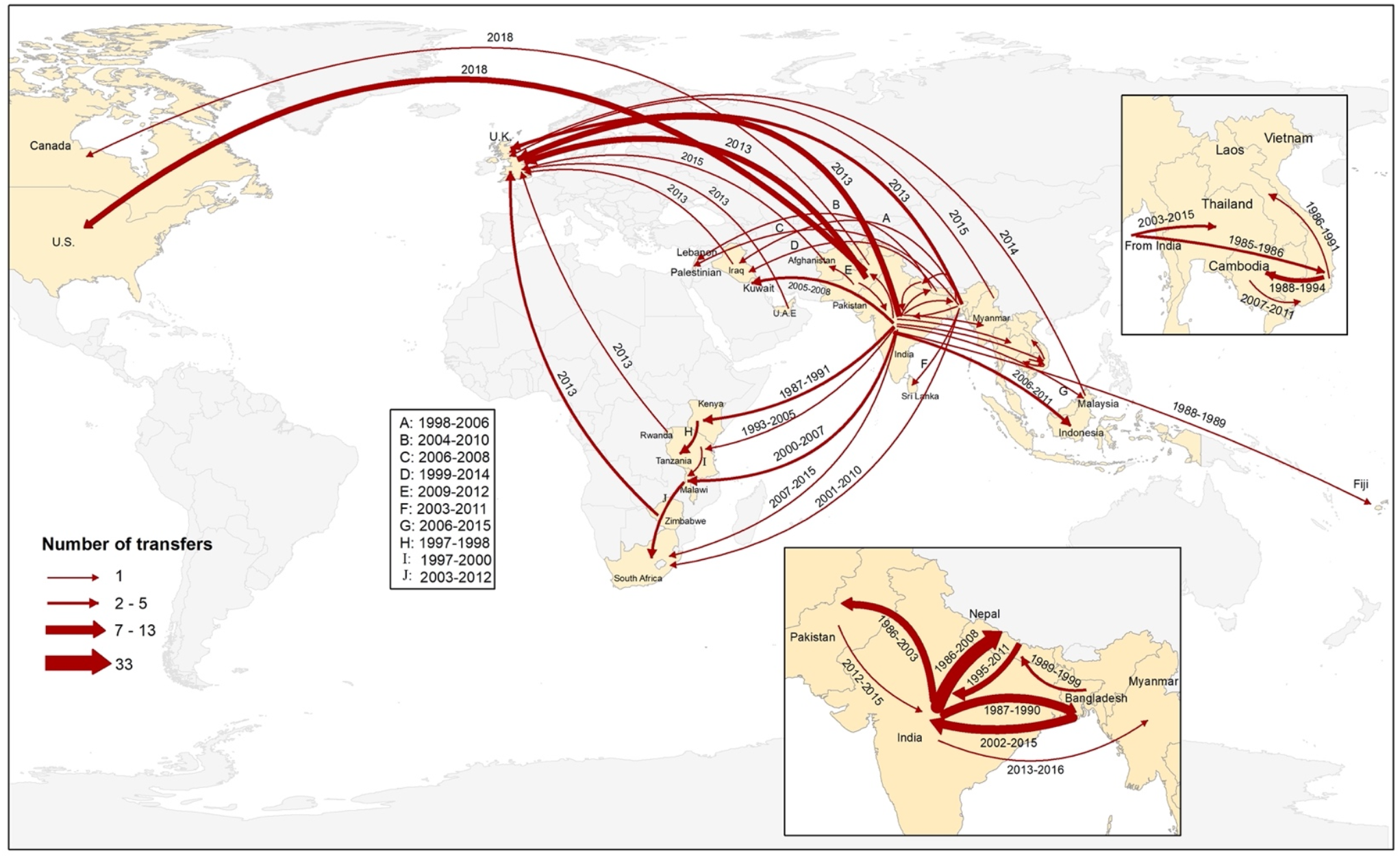
Geographical transfers within lineage 4.3.1 (H58) inferred from ancestral state reconstruction of the timed phylogenetic tree. The size of each arrow is scaled to the estimated number of transfers between the countries. Dates indicate the estimated first transfer between each pair of countries.

The major non-H58 clades also acquired AMR loci and spread within and from South Asia. Genotype 2.3.3 circulated predominantly in Bangladesh but spread to Pakistan and India within the past 30 years (**Figure S5**). QRDR mutations within genotype 2.3.3 have emerged independently on at least three occasions. Genotype 2.5, which may have circulated in India for >100 years (**Figure S6**), has been transferred to sub-Saharan Africa and Nepal multiple times, including two instances with strains containing QRDR mutations since 2015. Genotype 3.2.2 organisms originating from Bangladesh were observed in South Asia only. We observed a single instance of transfer from Bangladesh to Nepal and ongoing local expansion. In contrast, we found that these organisms have been regularly transferred from Bangladesh to India (**Figure S7**). Transfer events included at least four recent introductions of FQNS organisms between 2006 and 2017. The most recent common ancestor of genotype 3.3 was estimated to have been from India >200 years ago (**Figure S8**), but moved extensively across South Asia, establishing large subclades in Bangladesh and Nepal, before progressing to sub-Saharan Africa, the Middle East, and Southeast Asia. Genotype 3.3 organisms with QRDR mutations have moved from India to Nepal on multiple occasions.

Overall, our analysis identified evidence for 197 introduction events between countries, of which 138 were intracontinental and 59 were intercontinental (**Figure 5**). The most common international transmission events were within South Asia and from South Asia to Southeast Asia, East and Southern Africa. We estimated that resistance-conferring mutations to fluoroquinolones or azithromycin have independently emerged on at least 101 separate occasions within the last 30 years, mostly in South Asian countries (n = 94), and occasionally arising in Southeast Asia, Africa and South America. In addition, isolates carrying QRDR mutations were recently transferred between countries on at least 119 independent occasions.

**Figure 5.**
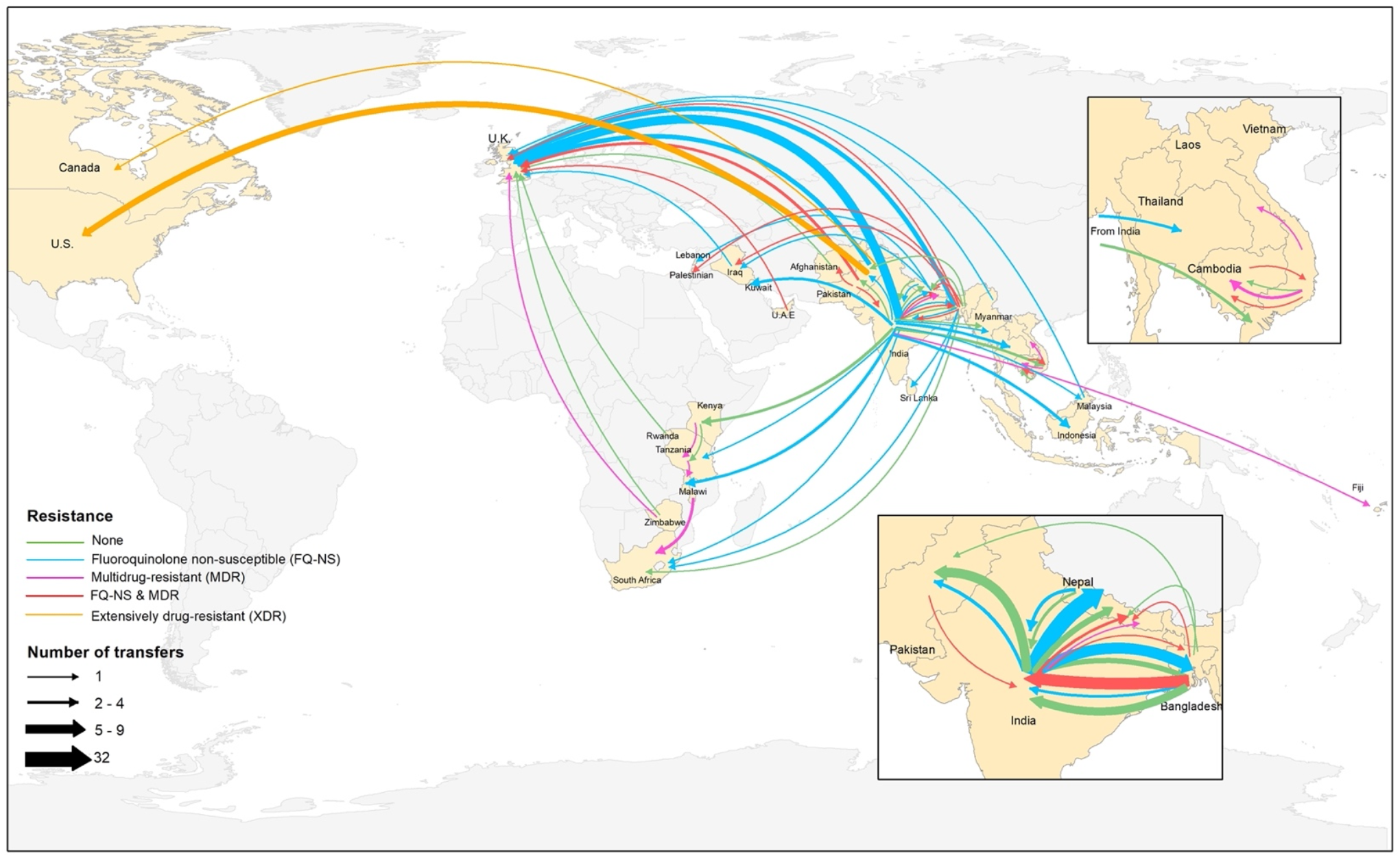
Major geographical transfers from 1990 onwards within the non-H58 and H58 lineages, inferred from the phylogenetic trees. The size of each arrow indicates the relative number of likely transfers between the countries. Arrow colors indicate antimicrobial resistance pattern.

## Discussion

This analysis of *S*. Typhi genome sequences reveals that the acquisition through plasmid acquisition or frequent, homoplastic mutations occurring across multiple lineages, has been accompanied by expansion and international spread of AMR *S*. Typhi clones. We identified numerous international and intercontinental transfers of *S*. Typhi over the past thirty years, with the majority associated with AMR phenotypes. Once introduced to a new setting, AMR *S*. Typhi become quickly fixed, as broadly exemplified with fluoroquinolone non-susceptible clades in multiple countries and XDR *S*. Typhi in Pakistan. This rapid emergence, spread and fixation of AMR in *S*. Typhi suggests that making decisions regarding TCV introduction via current AMR data may miss a critical window for prevention. Specifically, we found that South Asia continues to be an important hub for AMR generation in *S*. Typhi and that the clones emerging here regularly move internationally, underscoring the need for resources to support typhoid control in this region.

Our data are consistent with recent studies suggesting that MDR *S*. Typhi—strains resistant to the classical first line drugs— is now generally on the decline in South Asia (13,14). The decline of MDR *S*. Typhi in Asia has been accompanied by a decrease in the proportion of isolates carrying IncHI1 plasmids (except for Pakistan, where the MDR decline abruptly reversed amid the emergence of the XDR lineage). In our study, MDR was principally associated with H58 carrying chromosomally integrated AMR genes. The integration of AMR genes into the *S*. Typhi chromosome remains a concern, as it provides a mechanism for stable vertical transmission of MDR phenotypes (6,15). In contrast to South Asia, MDR typhoid associated with H58 and non-H58 isolates appears to be increasing in parts of Africa, with outbreaks being reported in the last decade (16,17).

QRDR mutations have independently arisen in all *S*. Typhi lineages due to sustained fluoroquinolone exposure. Nearly all of organisms containing QRDR mutations appear to have arisen in South Asia, and many have spread regionally and globally. Notably, our analysis revealed that highly fluoroquinolone resistant *S*. Typhi triple mutants have recently emerged in six different genotypes, including H58 lineage (4.3.1), lineage I (4.3.1.1), lineage II (4.3.1.2), and non-H58 isolates (3.3). Our phylogeography analysis suggests that these isolates most likely originated in India and disseminated to neighboring countries including Nepal and Pakistan.

The recent emergence and spread of resistance to third-generation cephalosporins and azithromycin further complicates typhoid fever treatment (8,9). Within three years of its first recognition, the XDR genotype (4.3.1.1.P1) became the dominant genotype circulating in Pakistan. The same sub-clade has been isolated from travelers returning from Pakistan to Canada, United Kingdom and United States (18,19). Recently, there have been reports of non-travel associated cases of 4.3.1.1.P1 XDR *S*. Typhi in the United States, suggestive of local transmission following importation (19). The XDR genotype has been typically associated with a composite MDR transposon inserted in the chromosome and by the acquisition of an IncY plasmid carrying *bla*_CTX-M-15_ and *qnrS1* (9). However, a recent study reported multiple integrations of *bla*_CTX-M-15_ from the IncY plasmid into the chromosome of XDR *S*. Typhi isolates (20). Chromosomal integration of *bla*_CTX-M-15_ may lessen its impact fitness cost, making it more likely that resistance will be maintained even in the absence of selection pressure in a comparable manner to the MDR gene cassette integration (15,20).

At present, all XDR *S*. Typhi strains identified have been susceptible to azithromycin and carbapenems (19). Concerningly, azithromycin-resistant *S*. Typhi have recently been reported in Bangladesh, India, Pakistan, Nepal and Singapore (10,21,22), arising from mutations in *acrB*. These mutations have arisen independently multiple times in distinct lineages (10). To date, XDR *S*. Typhi isolates containing mutations in *acrB* have not yet been identified. Such organisms would preclude effective treatment with established oral antimicrobials, which could lead to increased hospitalization rates, greater morbidity and mortality.

Our findings should be interpreted within the context of the limitations of the available data. While this analysis included the largest collection of novel *S*. Typhi genome sequences to date, there remains underrepresentation of sequences from several regions, including disproportionately small numbers from many countries in sub-Saharan Africa and Oceania where typhoid is endemic. Even in countries with more dense sampling, most isolates were derived from a small number of surveillance sites and may not be representative of the distribution of circulating strains. Because *S*. Typhi genomes only cover a fraction of all typhoid fever cases, phylogenies are highly incomplete; our estimates for AMR-conferring homoplastic mutations and international transfers represent lower bounds like may substantially underestimate their true frequency. These circumstances highlight the need for expanding genomic surveillance for enteric pathogens to provide a more comprehensive window into the emergence, expansion and spread of antimicrobial-resistant organisms.

The present study highlights the sustained emergence and geographic spread of AMR *S*. Typhi strains, with evidence of frequent international exportation. This observation underscores the importance of approaching typhoid fever control and antimicrobial resistance as a global rather than local problem. The recent emergence of XDR and azithromycin-resistant *S*. Typhi creates greater urgency for rapidly expanding prevention measures, including use of TCV and improvements to water and sanitation infrastructure, in typhoid-endemic countries. Such measures are needed in countries where AMR prevalence among *S*. Typhi isolates is currently high, but given the propensity for international spread, should not be restricted to such setting.

## Methods

### Bacterial isolates

This study included *S*. Typhi isolates obtained from the Surveillance for Enteric Fever in Asia Project (SEAP; Bangladesh, Nepal, and Pakistan; 2016-2019), Etiologies of Acute Febrile Illness Study (Nepal; 2014-2016), and Surveillance for Enteric Fever in India Project (SEFI; 2017-2019. The study methodologies have been previously described (23–25). In brief, participants for these studies included individuals of all ages presenting to study site facilities with febrile illness.

These included 5 facilities in Dhaka, Bangladesh, 18 facilities across 16 cities in India, 11 facilities across 3 cities in Nepal and two hospitals and a university laboratory network in Karachi, Pakistan. Across these studies, there were a total of 9,945 blood culture-confirmed typhoid cases. From these, we selected a country-stratified sample of 3,489 *S*. Typhi isolates for sequencing. Details on the sampling for each country are available in the Appendix.

### Whole-genome sequencing

Whole-genome sequencing (WGS) was performed at the Wellcome Trust Sanger Institute using the Illumina Hiseq2500 platform (Illumina, San Diego, CA, USA) to generate paired-end reads of 100–150 bp in length, and at a commercial service in Bangalore and at the Wellcome Trust Research Laboratory in the Christian Medical College using the Illumina Miseq platform (Illumina, San Diego, CA, USA). Sequence data quality was checked using FastQC v0.11.9 to remove low quality reads (26). We summarized all quality indicators using MultiQC v1.7 (27). Species identification was confirmed with Kraken2 (28), and the *Salmonella in silico* Typing Resource (SISTR) was used for WGS-based serotyping (29). Short Read Sequence Typing for Bacterial Pathogens (SRST2) (30) was used to map known alleles and identify MLSTs directly from reads according to the *Salmonella enterica* MLST scheme (https://pubmlst.org/salmonella/).

### Mapping and SNP analysis

Paired-end Illumina reads were mapped to the *S*. Typhi CT18 (accession no. AL513382) reference chromosome sequence using RedDog mapping pipeline v1beta.11 (https://github.com/katholt/reddog). RedDog uses Bowtie2 v2.4.1 (31) to map reads to the reference genome, and SAMtools v1.10 (32) to identify SNPs that have a phred quality score above 30, and to filter out those SNPs supported by less than five reads, or with 2.5x the average read depth that represent putative repeated sequences, or those that have ambiguous base calls. For each SNP that passes these criteria in any one isolate, consensus base calls for the SNP locus were extracted from all genomes mapped, with those having phred quality scores under 20 being treated as unknown alleles and represented with a gap character.

Chromosomal SNPs with confident homozygous calls (phred score above 20) in >95% of the genomes mapped (representing a ‘soft’ core genome) were concatenated to form an alignment of alleles using the RedDog python script parseSNPtable.py with parameters -m cons, aln and –c 0.95 and SNPs called in prophage regions and repetitive sequences (354 kb; ∼7.4% of bases) in the CT18 reference chromosome, as defined previously (33) were excluded to form an alignment of 14,901 variant sites. SNPs occurring in recombinant regions were detected by Gubbins v2.4.1 (34) and excluded resulting in a final alignment of 11,978 chromosomal SNPs. The SNP data were used to assign all isolates to previously defined genotypes according to an extended *S*. Typhi genotyping framework using the GenoTyphi python script (https://github.com/katholt/genotyphi) (33). To provide global context, additional *S*. Typhi genomes (9,12,17,33,35–41) were subjected to both SNP calling, recombination filtering, and genotyping as described above, resulting in an alignment of 28,897chromosomal SNPs.

### Phylogenetic analyses

Maximum likelihood (ML) phylogenetic trees were inferred from the chromosomal SNP alignments using RAxML v8.2.10 (42) (command raxmlHPC-PTHREADS). A generalized time-reversible model and a Gamma distribution was used to model site-specific rate variation (GTR+ Γ substitution model; GTRGAMMA in RAxML) with 100 bootstrap pseudoreplicates used to assess branch support for the ML phylogeny. We selected the single tree with the highest likelihood score as the best tree. The resulting phylogenies were visualized and annotated using the iTOL v5 online version (43).

### Temporal and phylogeographic analysis

To investigate *dates* of emergence and geographical transfers, we inferred timed phylogenies using globally and temporally representative samples. First, we used TempEst v1.5 to assess temporal structure by conducting a regression of the root-to-tip branch distances of the tree as a function of the sampling time (44), which was confirmed by a clustered permutation test using *BactDating* (45). For the non-H58 isolates, we estimated the best-fit models, tree topology, evolutionary rates, and phylogeography by using a Bayesian Markov chain Monte Carlo (MCMC) method with the software package BEAST2 v2.6.2 (46). Separate trees were fit for each of the most common non-H58 lineages (2.3.3, 2.5, 3.2.2, and 3.3). Isolates from each lineage were selected based on temporal, geographic and phylogenetic diversity, as described in detail in the Appendix.

For the BEAST analysis, a GTR+Γ substitution model was selected, and the sampling times (tip dates) were defined as the year of isolation to calibrate the molecular clock. We tested support for a strict clock for each lineage using the relaxed clock test in *treedater*, and the strict clock was rejected in each instance (47). We therefore constructed time-phylogenies using coalescent exponential population priors with a relaxed clock (uncorrelated lognormal distribution) (6,17). Three independent runs were performed to ensure convergence, and were combined with LogCombiner, following removal of the first 10 million steps from each as burn-in. The effective sample sizes (ESSs) of the parameters were estimated to be >200 for all independent runs of the analysis. The trees were summarized in a maximum clade credibility (MCC) target tree using the Tree Annotator program v2.4.7. The time of the most recent common ancestor (tMRCA) estimates were calculated as the years before the most recent sampling dates. Phylogeographical reconstruction was obtained by the continuous-time Markov Chain process over discrete sampling locations implemented in BEAST. The final trees were visualized using FigTree v1.4 (http://tree.bio.ed.ac.uk/software). For H58, we had 4,761 isolates, which precluded temporal and phylogeographic analysis using BEAST due to computational constraints. To avoid significant down-sampling of isolates, we used the *treedater* R package (47) with an uncorrelated, relaxed molecular lock to estimate the timed phylogeny for H58, which yielded a tMRCA matching a root-to-tip based analysis using *BactDating* and consistent with previously published estimates (6). We included all H58 sequences available in our collection to improve accuracy of the timed phylogeny and phylogeographic analysis. We reconstructed the ancestral state of nodes using the maximum parsimony approach with the *Phangorn* R package, considering important events with a location probability of >0.5 between connected nodes. For visualization purposes, we selected a smaller subset of sequences to depict in a dated phylogenetic tree. For all phylogeographic analyses, we considered a geographic transfer when the most probable location between two connected nodes (or between a node and a tip) differed, and we considered the time window of transfer as the date range between the nodes (or between the node and tip). The geospatial transmissions of lineage strains from the phylogeographic reconstructions were analyzed and visualized using ArcMap 10.7.1 (https://desktop.arcgis.com/en/arcmap/).

### Non-parametric phylodynamic inference of effective population size

To evaluate the historical effective population size for H58 lineage strains, we used the time-stamped H58 tree to estimate the effective population size through time using the *skygrowth* package (https://github.com/mrc-ide/skygrowth). We pruned the timed phylogenies by country and antimicrobial resistance pattern to compare the effective population sizes of antimicrobial resistant and sensitive populations.

### AMR associated genes detection and plasmid replicon analysis

ARIBA (Antimicrobial Resistance Identifier by Assembly) v2.10.0 and CARD database v1.1.8 (https://card.mcmaster.ca/home) were used to investigate antimicrobial resistance gene content. Point mutations in the quinolone resistance determining region (QRDR) of the DNA-gyrase *gyrA/B* and topoisomerase-IV *parC/E* genes, associated with reduced susceptibility to fluoroquinolones and quinolone resistance genes (*qnrS*) were also detected using ARIBA. Isolates were defined as being MDR if resistance genes were detected by ARIBA in the beta-lactam, trimethoprim/sulphonamide and chloramphenicol classes. Plasmid replicons were identified using ARIBA and the PlasmidFinder database (30).

## Supporting information

Summary of the 3,489 samples sequenced in this study

Summary of the 4,169 sequences previously published included in this study

## Data Availability

Illumina sequence data was submitted to the European Nucleotide Archive. Sequence data from 4,169 S. Typhi strains from previous studies were also included for global context raw sequence data for these isolates are available in European Nucleotide Archive. Details and individual accession numbers of sequence data included in our analysis have been included in Tables S1 and S2.

## Data availability

Illumina sequence data was submitted to the European Nucleotide Archive. Sequence data from 4,169 *S*. Typhi strains from previous studies were also included for global context raw sequence data for these isolates are available in European Nucleotide Archive. Details and individual accession numbers of sequence data included in our analysis have been included in Tables S1 and S2.

## Ethics statement

Ethical approval for the parent studies were obtained from the Bangladesh Institute of Child Health Ethical Review Committee, Christian Medical College Institutional Review Board, Nepal Health Research Council, Aga Khan University Hospital Ethics Committee and Pakistan National Ethics Committee, Stanford University Institutional Review Board, and U.S. Centers for Disease Control and Prevention. Informed written consent and clinical information were taken from adult participants and legal guardians of child participants.

## Acknowledgements

This work was supported by a grant from the Bill and Melinda Gates Foundation (grant number INV-008335).

## Notes

### Competing Interest Statement

The authors have declared no competing interest.

