## Supplemental material for "The international and intercontinental spread and expansion of antimicrobial-resistant *Salmonella* Typhi"

**Supplementary Figure 1.** Frequency and distribution of genotypes of contemporary *Salmonella* Typhi isolates included in our study (2014 to 2019). All the genotypes present in in less than three samples are not shown. 2* includes 2, 2.0.1, 2.0.2, 2.1.7, 2.2.1, 2.2.2, 2.2.4, 2.3.4 and 2.4 genotypes. 3* includes 3, 3.0.1, 3.1.2 and 3.2.1 genotypes.

**Supplementary Figure 2.** Maximum likelihood tree of 3,489 contemporary *S*. Typhi isolates from South Asia. Inner ring indicates the genotypes. The second ring indicates QRDR mutations profiles. Third ring indicates the presence of *acrB* mutations associated with azithromycin-resistant isolates. Fourth ring indicates MDR or XDR status, and the outer ring indicates country of isolation. The scale bar indicates nucleotide substitutions per site.

**Supplementary Figure 3.** Antimicrobial resistance trends of *Salmonella* Typhi isolates. **(a)** Proportion and temporal distribution of multidrug-resistant (MDR) *S*. Typhi isolates from our global collection. Isolates were considered MDR if they contained genes conferring resistance to chloramphenicol (*catA1*), trimethoprim-sulfamethoxazole (*dfrA7*, *sul1*, or *sul2*) and ampicillin ($bla_{\text{TEM-1}}$). Shading indicates missing data. Bars indicate temporal distribution of isolates. **(b)** Proportion and temporal distribution of Fluoroquinolone non-susceptible (FQ-NS) *S*. Typhi isolates from our global collection. We classified isolates as FQ-NS if they carried quinolone resistance genes (*qnrS*), and/or point mutations in the quinolone resistance determining region (QRDR) of the DNA-gyrase *gyrA/B* and topoisomerase-IV *parC/E***.**

**Supplementary Figure 4.** Frequency of chromosomal point mutations in the quinolone resistance-determining region (QRDR) per isolate over time.

**Supplementary Figure 5**. Phylogeography and global expansion of *S*. Typhi lineage 2.3.3. **(a)** Maximum clade credibility tree (reconstructed using BEAST2) of genotype 2.3.3 *S*. Typhi isolates.

The branches are in time scale in years and are colored according to the location of the most probable ancestor of descendant nodes. The scale bar indicates nucleotide substitutions per site. **(b)** Geographical transfers within the 2.3.3 lineage, inferred from the phylogenetic tree. The size of each arrow indicates the relative number of likely transfers between the countries.

**Supplementary Figure 6**. Phylogeography and global expansion of *S.* Typhi lineage 2.5. **(a)** Maximum clade credibility tree (reconstructed using BEAST2) of genotype 2.5 *S*. Typhi isolates. The branches are in time scale in years and are colored according to the location of the most probable ancestor of descendant nodes. Branches are displayed as dashed lines when the posterior probability values were below 0.5. The scale bar indicates nucleotide substitutions per site. **(b)** Geographical transfers within the 2.5 lineage, inferred from the phylogenetic tree. The size of each arrow indicates the relative number of likely transfers between the countries.

**Supplementary Figure 7.** Phylogeography and global expansion of lineage 3.2.2. **(a)** Maximum clade credibility tree (reconstructed using BEAST2) of genotype 3.2.2. *S*. Typhi isolates. The branches are in time scale in years and are colored according to the location of the most probable ancestor of descendant nodes. Branches are displayed as dashed lines when the posterior probability values were below 0.5. The scale bar indicates nucleotide substitutions per site. **(b)** Geographical transfers within the 3.2.2 lineage, inferred from the phylogenetic tree. The size of each arrow indicates the relative number of likely transfers between the countries.

**Supplementary Figure 8.** Phylogeography and global expansion of *S.* Typhi lineage 3.3. **(a)** Maximum clade credibility tree (reconstructed using BEAST2) of genotype 3.3 *S*. Typhi isolates. The branches are in time scale in years and are colored according to the location of the most probable ancestor of descendant nodes. Branches are displayed as dashed lines when the posterior probability values were below 0.5. The scale bar indicates nucleotide substitutions per site. **(b)** Geographical transfers within the lineage 3.3 inferred from the phylogenetic tree. The size of each arrow indicates the relative number of likely transfers between the countries.

**Supplementary Table 1.** Summary of the 3,489 samples sequenced in this study.

**Supplementary Table 2.** Summary of the 4,169 sequences previously published included in this study.

**Supplementary Table 3.** Frequency of QRDR mutation patterns present among 7,658 *S*. Typhi isolates from the global collection.

**Supplementary Table 4.** Estimated times of the most recent common ancestors (tMRCAs) of the main clades, with calendar years and most probable locations, from discrete phylogeography of the *S*. Typhi isolates.

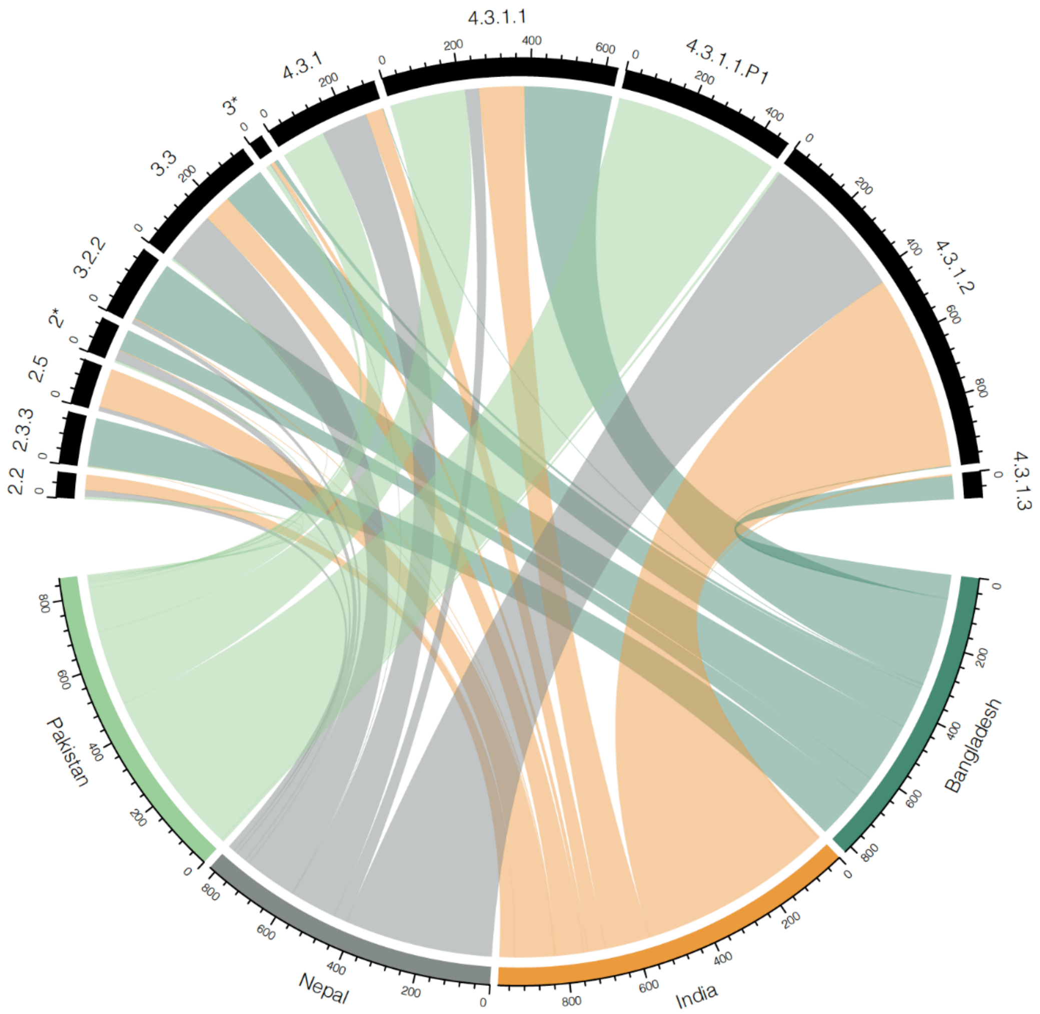


**Supplementary Figure 1.** Frequency and distribution of genotypes of contemporary *Salmonella* Typhi isolates included in our study (2014 to 2019). All the genotypes present in in less than three samples are not shown. 2* includes 2, 2.0.1, 2.0.2, 2.1.7, 2.2.1, 2.2.2, 2.2.4, 2.3.4 and 2.4 genotypes. 3* includes 3, 3.0.1, 3.1.2 and 3.2.1 genotypes.

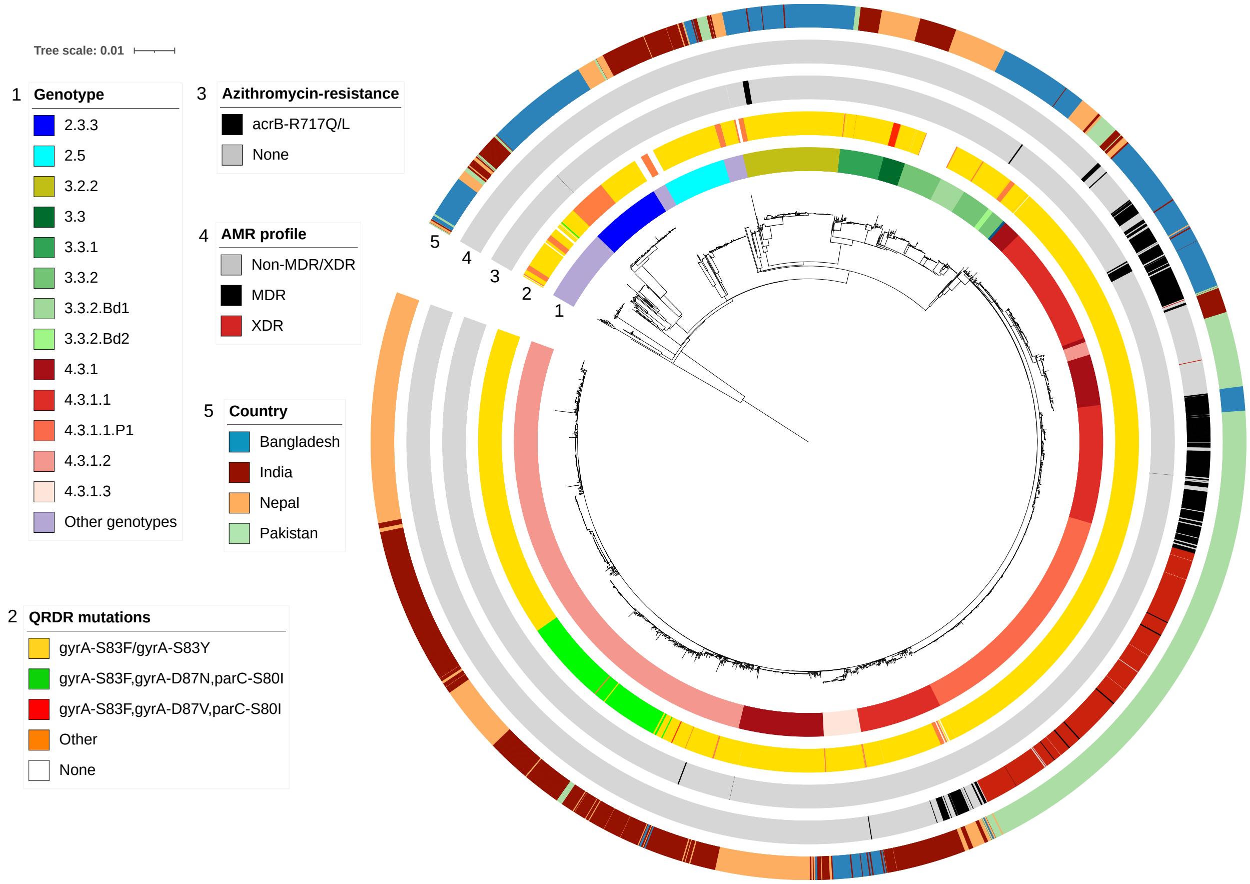


**Supplementary Figure 2.** Maximum likelihood tree of 3,489 contemporary *S.* Typhi isolates from South Asia. Inner ring indicates the genotypes. The second ring indicates QRDR mutations profiles. Third ring indicates the presence of *acrB* mutations associated with azithromycin-resistant isolates. Fourth ring indicates MDR or XDR status, and the outer ring indicates country of isolation. The scale bar indicates nucleotide substitutions per site.

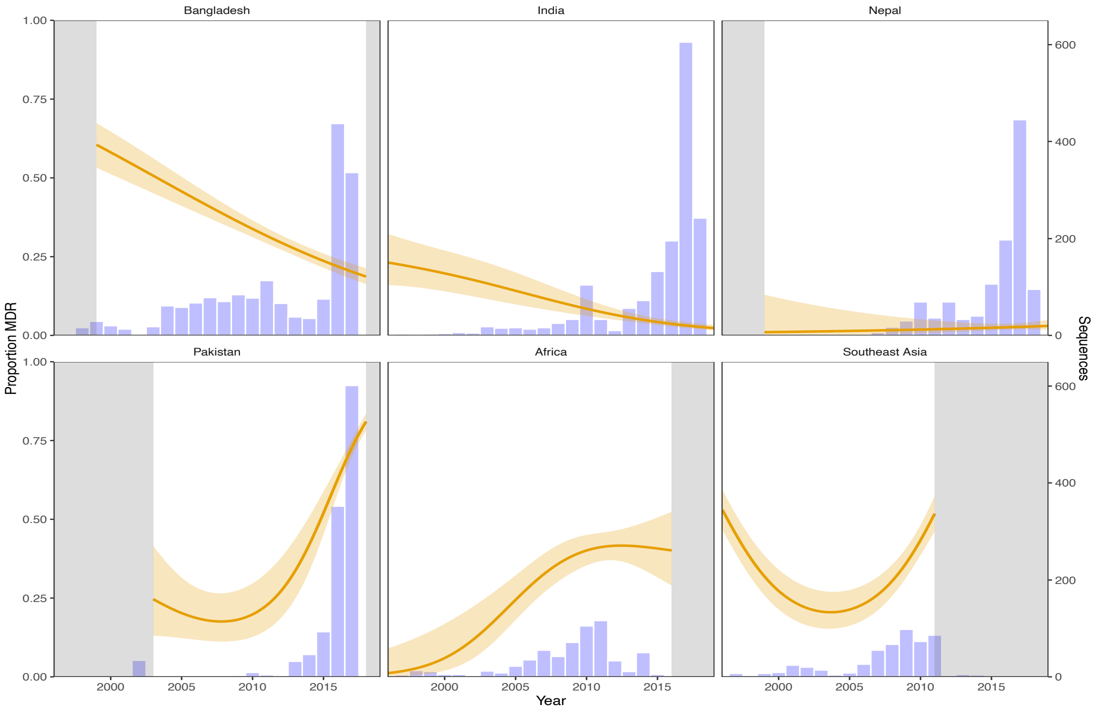


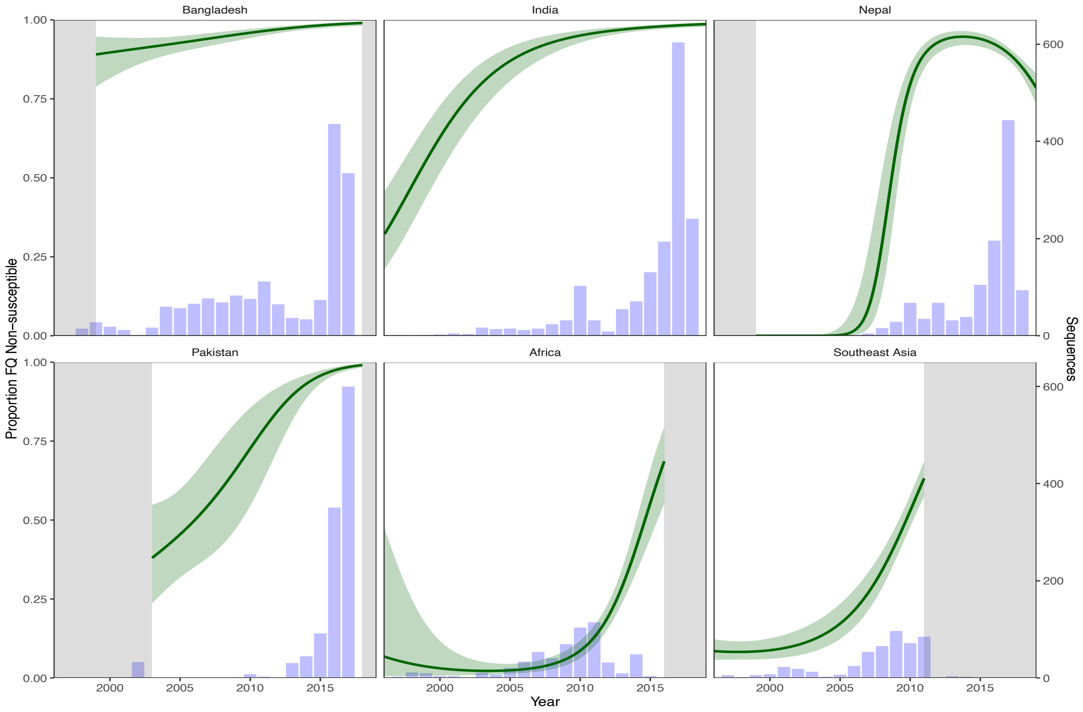


**Supplementary Figure 3.** Antimicrobial resistance trends of *Salmonella* Typhi isolates. **(a)** Proportion and temporal distribution of multidrug-resistant (MDR) *S*. Typhi isolates from our global collection. Isolates were considered MDR if they contained genes conferring resistance to chloramphenicol (*catA1*), trimethoprim-sulfamethoxazole (*dfrA7*, *sul1*, or *sul2*) and ampicillin (*bla*$_{TEM-1}$). Shading indicates missing data. Bars indicate temporal distribution of isolates. **(b)** Proportion and temporal distribution of Fluoroquinolone non-susceptible (FQ-NS) *S*. Typhi isolates from our global collection. We classified isolates as FQ-NS if they carried quinolone resistance genes (*qnrS*), and/or point mutations in the quinolone resistance determining region (QRDR) of the DNA-gyrase *gyrA/B* and topoisomerase-IV *parC/E*.

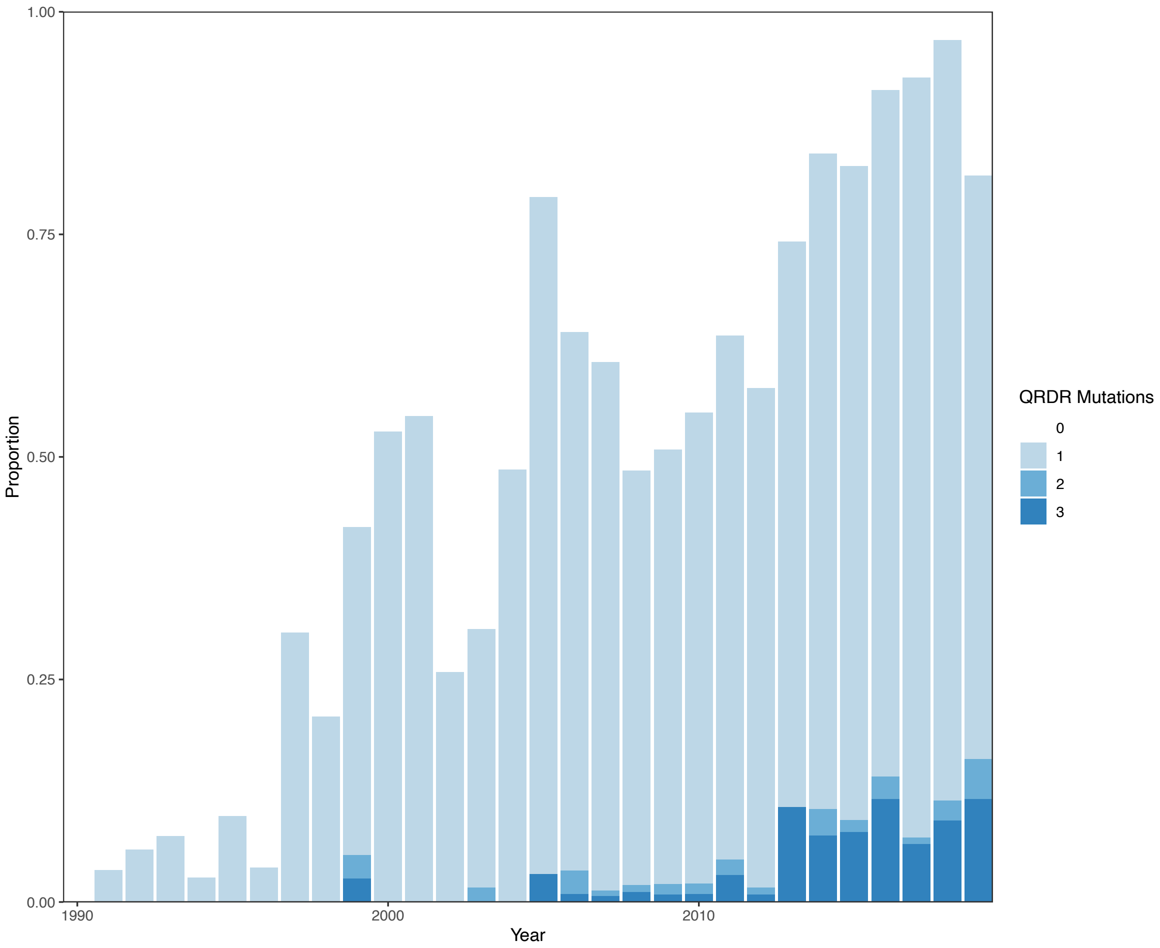


**Supplementary Figure 4.** Frequency of chromosomal point mutations in the quinolone resistance-determining region (QRDR) per isolate over time.

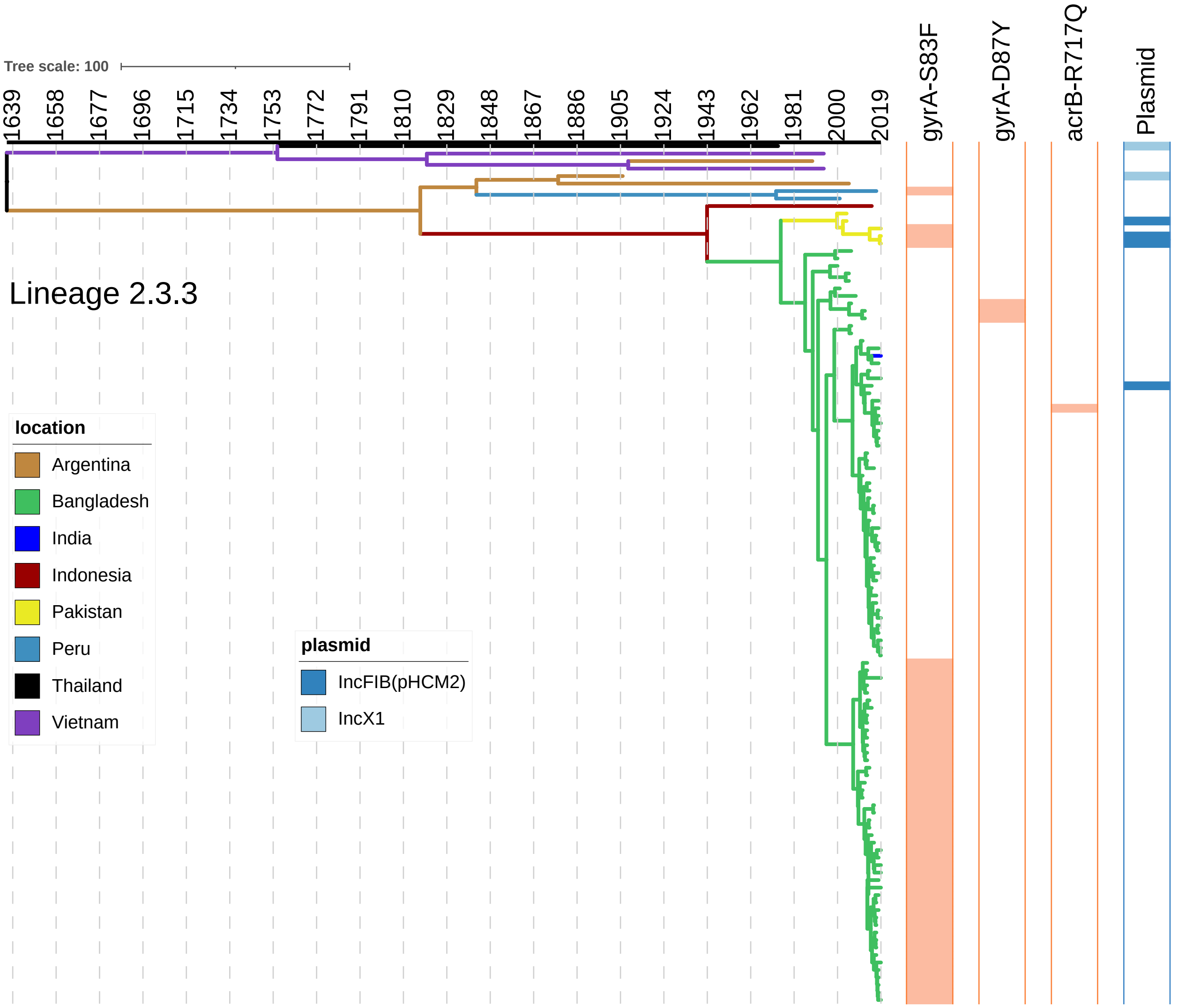


**Supplementary Figure 5**. Phylogeography and global expansion of *S.* Typhi lineage 2.3.3. **(a)** Maximum clade credibility tree (reconstructed using BEAST2) of genotype 2.3.3 *S*. Typhi isolates. The branches are in time scale in years and are colored according to the location of the most probable ancestor of descendant nodes. The scale bar indicates nucleotide substitutions per site.

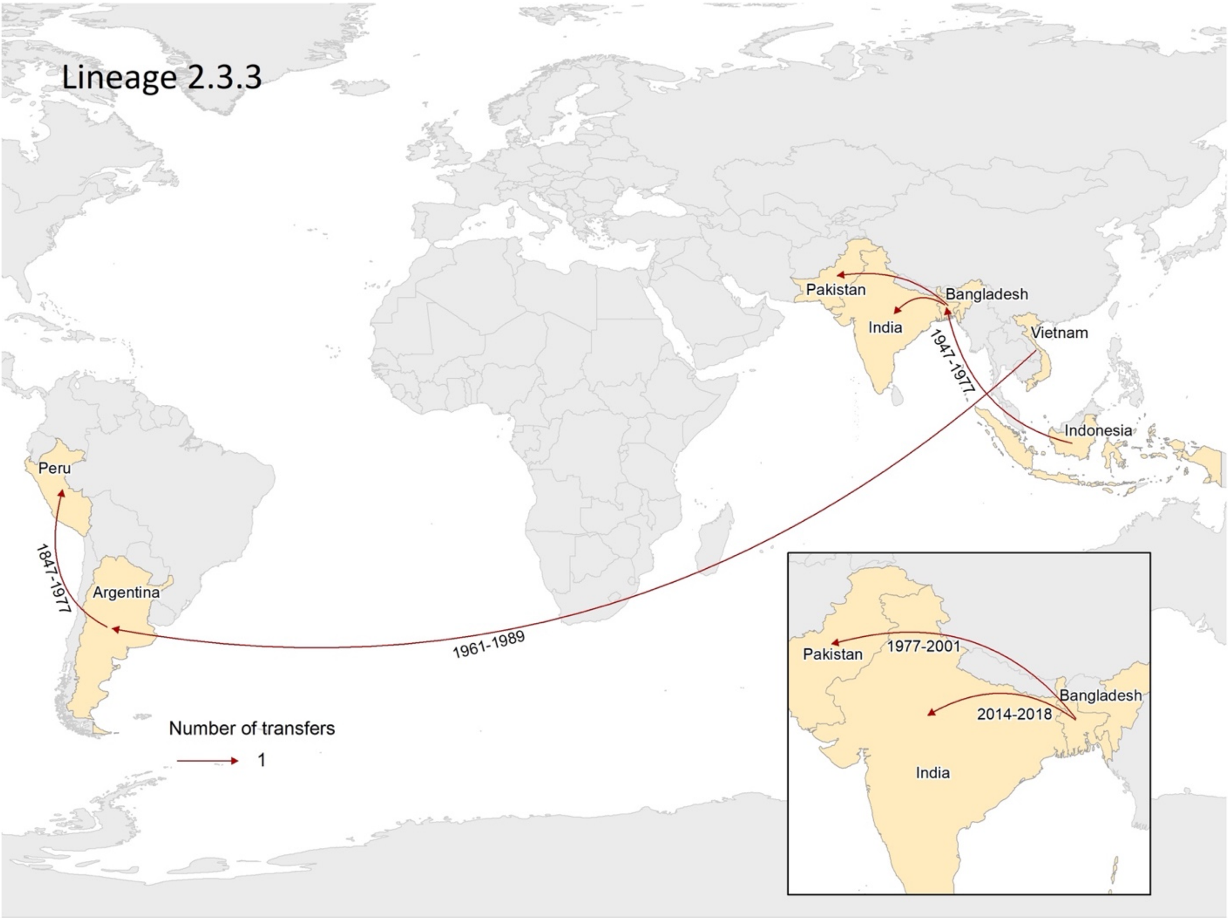


**(b)** Geographical transfers within the 2.3.3 lineage, inferred from the phylogenetic tree. The size of each arrow indicates the relative number of likely transfers between the countries.

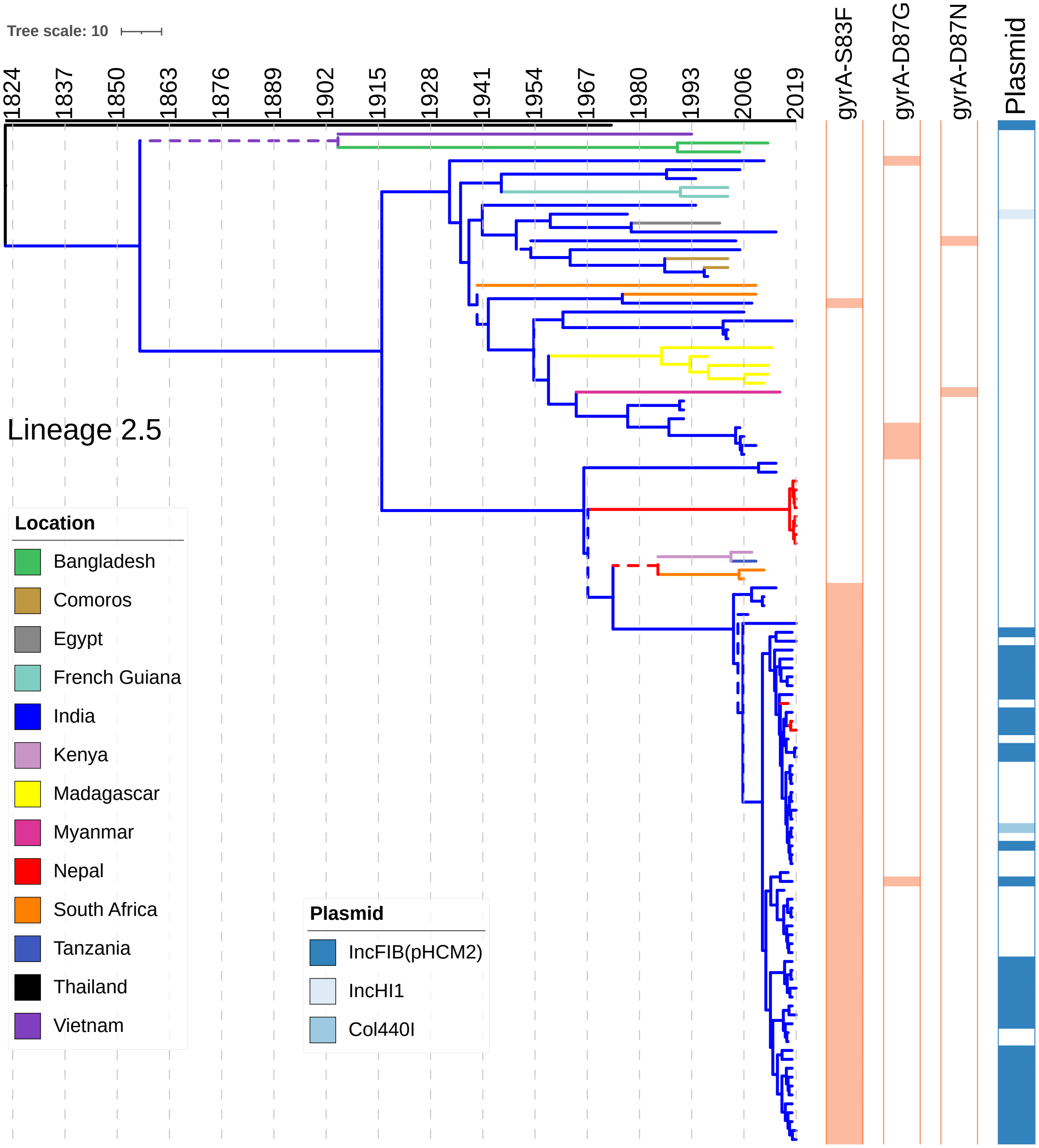


**Supplementary Figure 6**. Phylogeography and global expansion of *S.* Typhi lineage 2.5. **(a)** Maximum clade credibility tree (reconstructed using BEAST2) of genotype 2.5 *S.* Typhi isolates. The branches are in time scale in years and are colored according to the location of the most probable ancestor of descendant nodes. Branches are displayed as dashed lines when the posterior probability values were below 0.5. The scale bar indicates nucleotide substitutions per site.

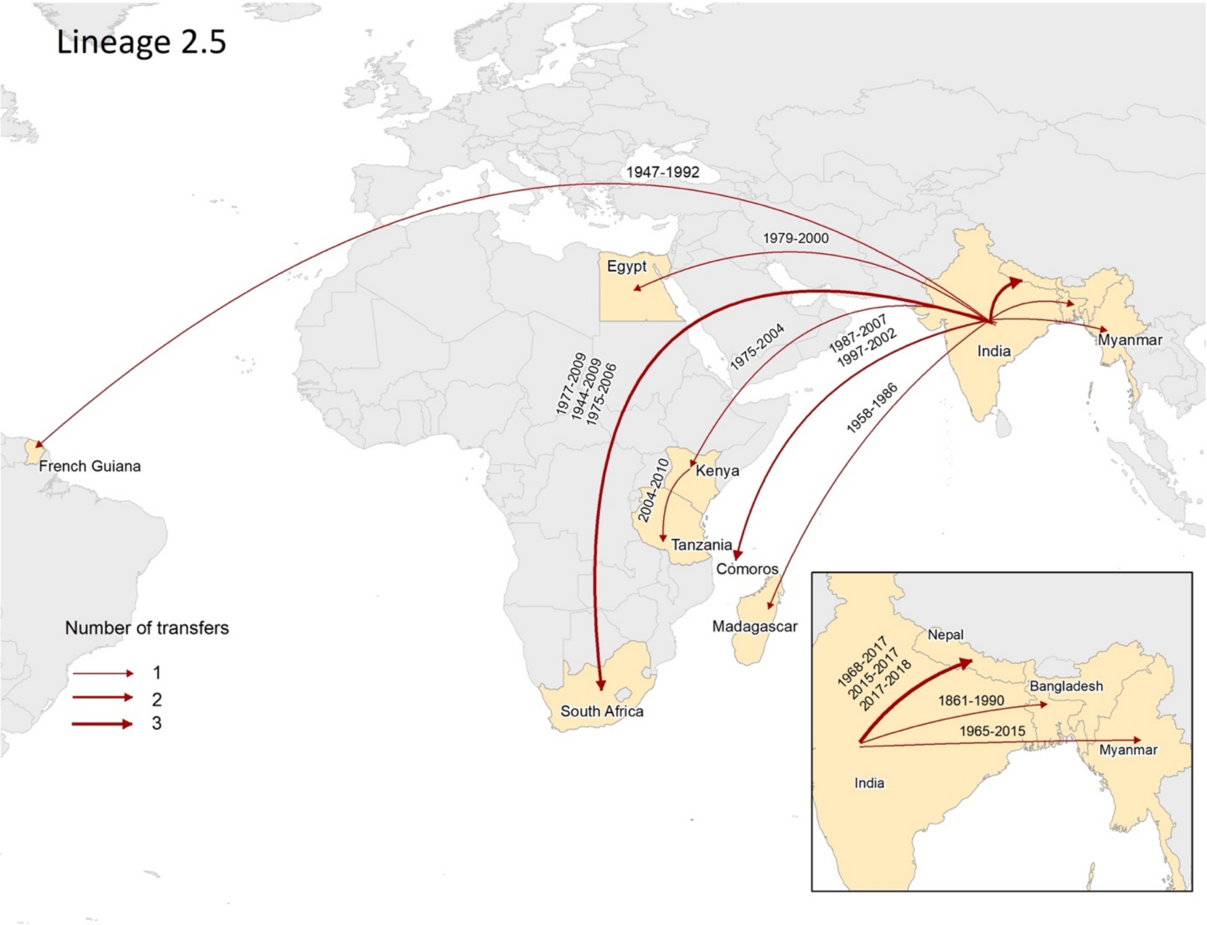


**(b)** Geographical transfers within the 2.5 lineage, inferred from the phylogenetic tree. The size of each arrow indicates the relative number of likely transfers between the countries.

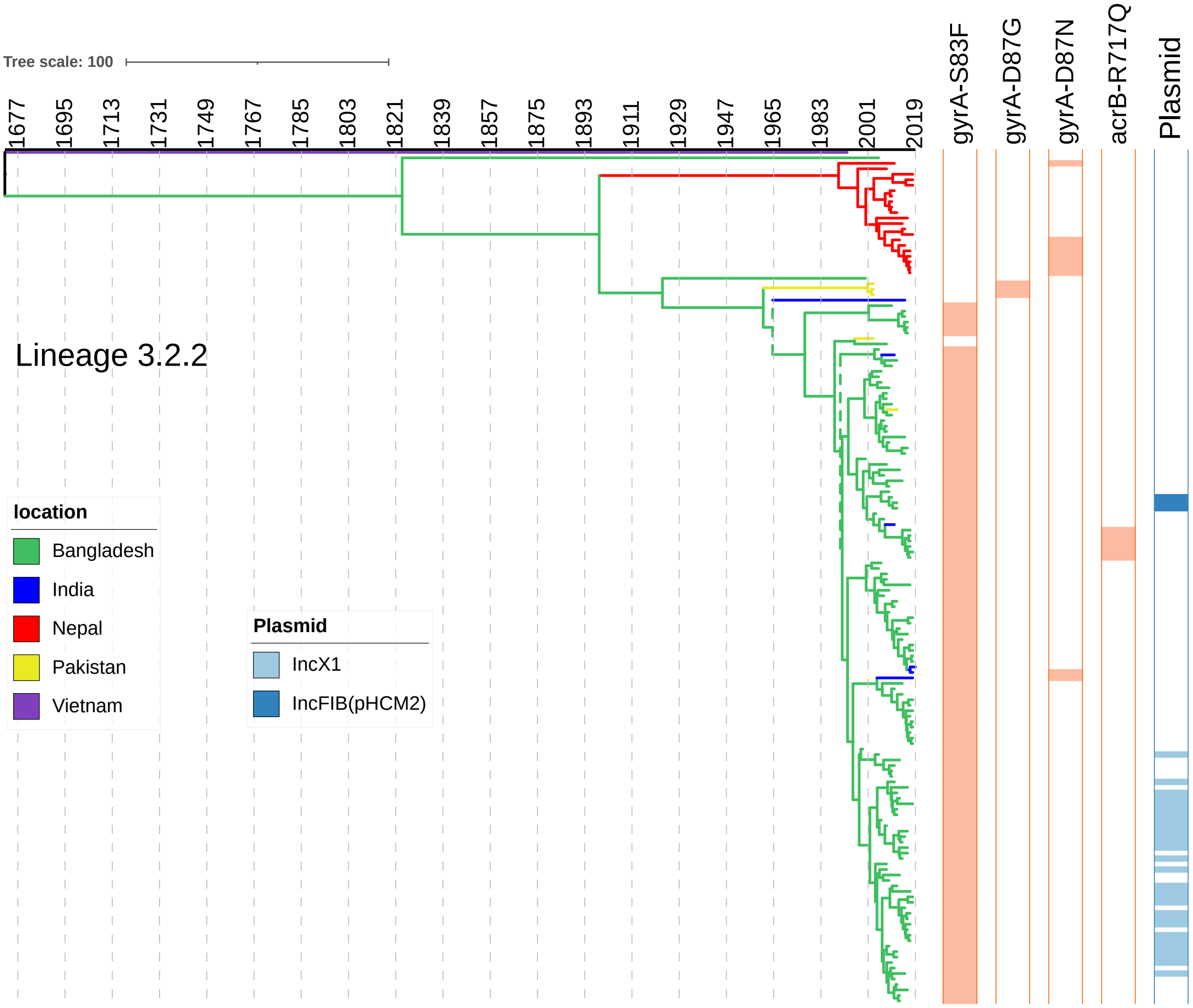


**Supplementary Figure 7.** Phylogeography and global expansion of lineage 3.2.2. **(a)** Maximum clade credibility tree (reconstructed using BEAST2) of genotype 3.2.2. *S.* Typhi isolates. The branches are in time scale in years and are colored according to the location of the most probable ancestor of descendant nodes. Branches are displayed as dashed lines when the posterior probability values were below 0.5. The scale bar indicates nucleotide substitutions per site.

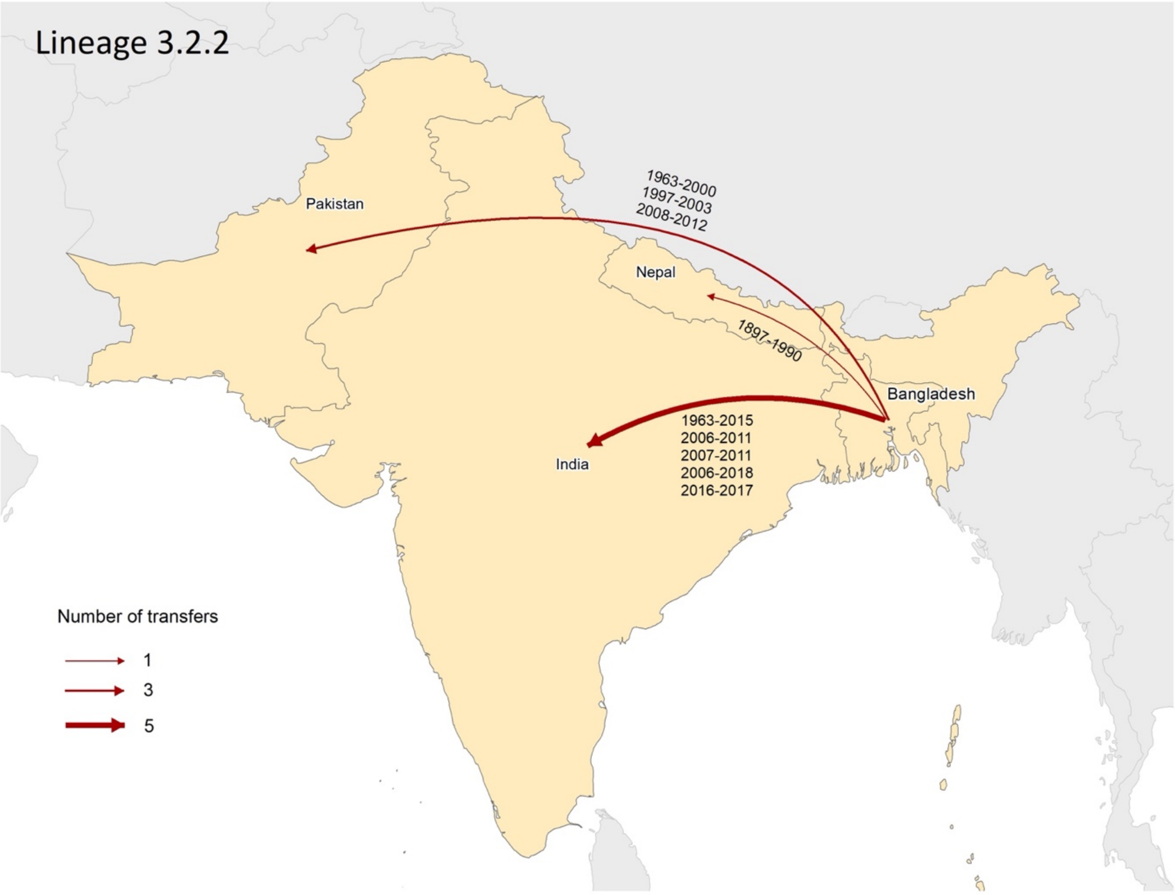


**(b)** Geographical transfers within the 3.2.2 lineage, inferred from the phylogenetic tree. The size of each arrow indicates the relative number of likely transfers between the countries.

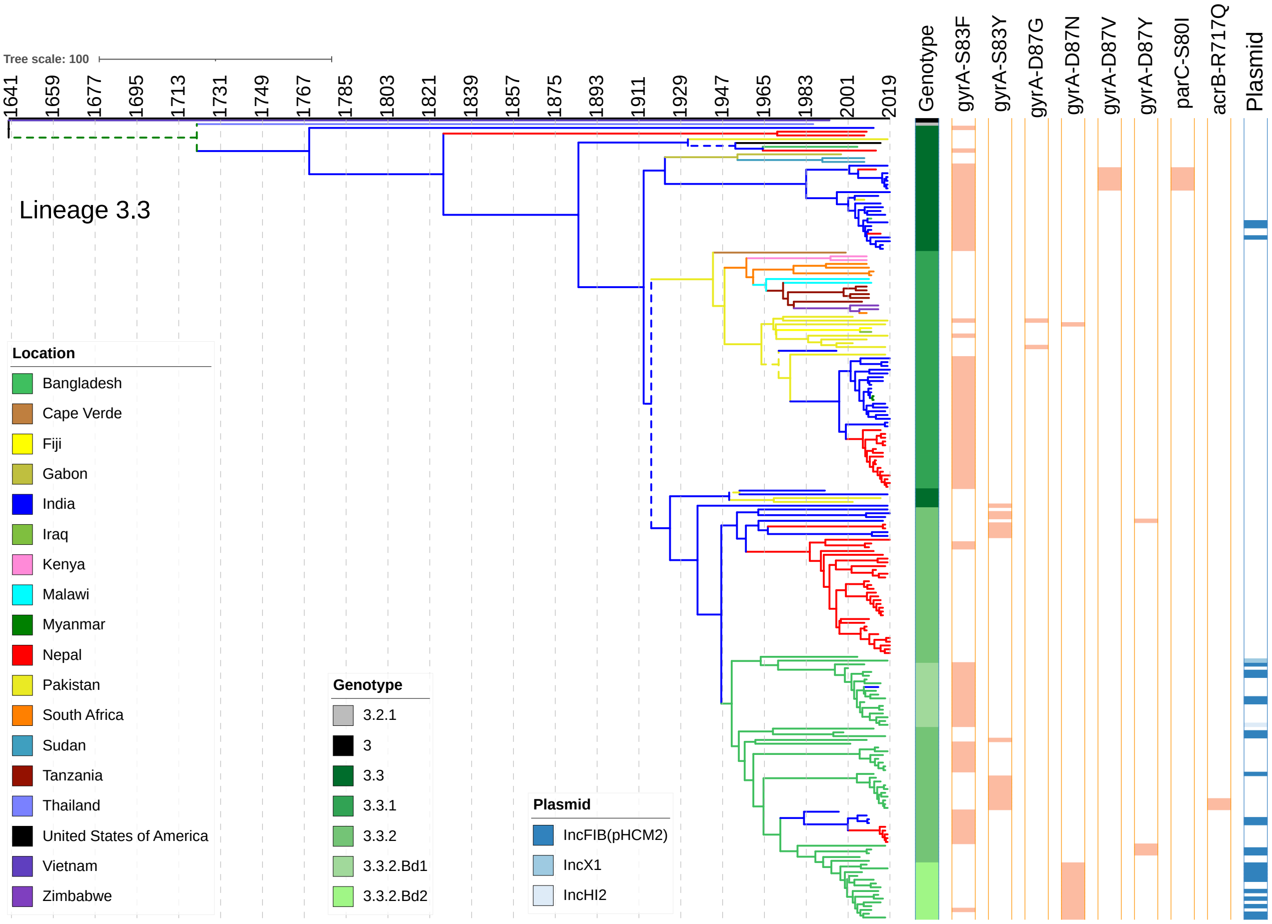


**Supplementary Figure 8.** Phylogeography and global expansion of *S.* Typhi lineage 3.3. **(a)** Maximum clade credibility tree (reconstructed using BEAST2) of genotype 3.3 *S.* Typhi isolates. The branches are in time scale in years and are colored according to the location of the most probable ancestor of descendant nodes. Branches are displayed as dashed lines when the posterior probability values were below 0.5. The scale bar indicates nucleotide substitutions per site.

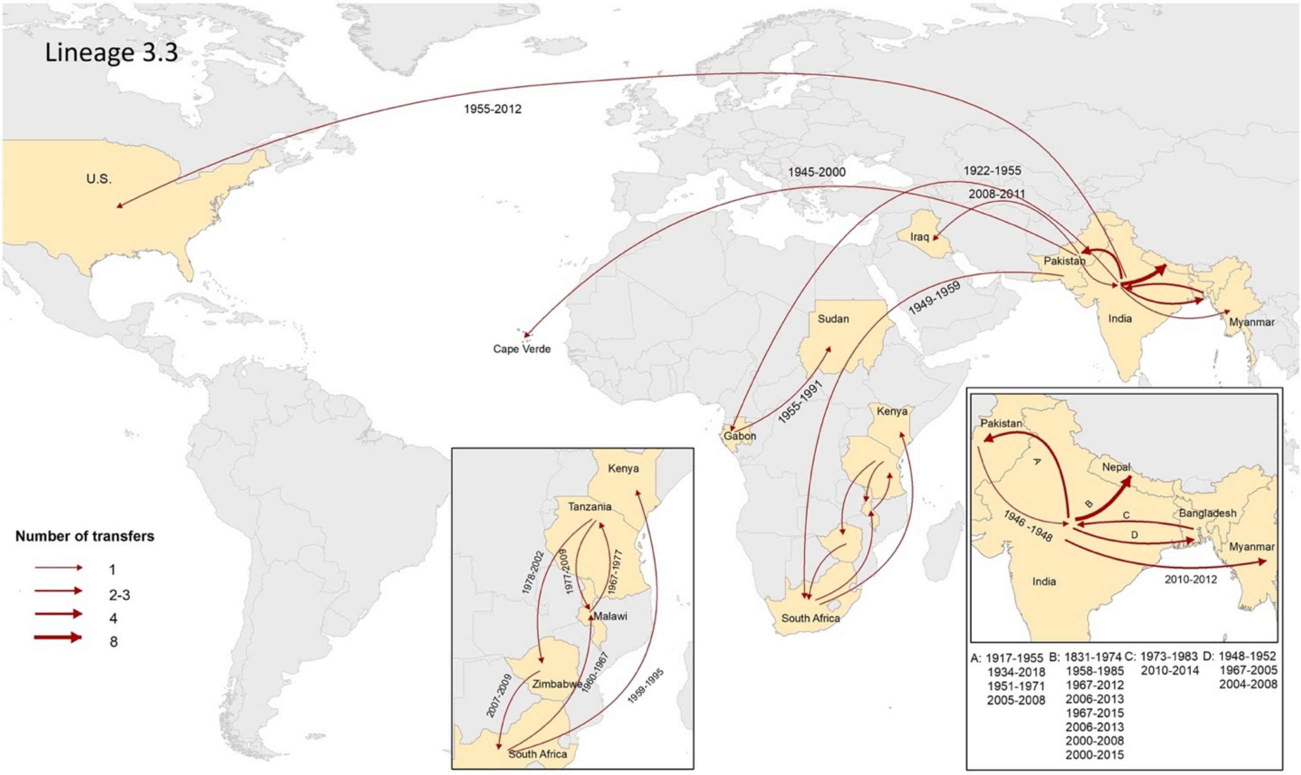


**(b)** Geographical transfers within the lineage 3.3 inferred from the phylogenetic tree. The size of each arrow indicates the relative number of likely transfers between the countries.

**Supplementary Table 3.** Frequency of QRDR mutation patterns present among 7,658 *S.* Typhi isolates from the global collection.

| **Mutation profile** | | **Number of isolates n = 5613 (%)** |
|---|---|---|
| **Single mutant** | | |
| | gyrA-S83F | 3950 (70.39) |
| | gyrA-S83Y | 927 (16.52) |
| | gyrA-D87N | 126 (2.24) |
| | gyrA-D87G | 32 (0.57) |
| | gyrA-D87Y | 17 (0.30) |
| | parC-S80I | 4 (0.07) |
| **Total** | | 5057 (90.09) |
| **Double mutant** | | |
| | gyrA-S83F, parC-S80I | 57 (1.02) |
| | gyrA-S83F, parC-E84G | 38 (0.68) |
| | gyrA-S83F, parC-E84K | 13 (0.23) |
| | gyrA-D87N, gyrA-S83F | 4 (0.07) |
| | gyrA-S83Y, parC-S80R | 4 (0.07) |
| | gyrA-D87G, gyrA-S83F | 2 (0.04) |
| | gyrA-S83F, parC-S80R | 1 (0.02) |
| **Total** | | 119 (2.12) |
| **Triple mutant** | | |
| | gyrA-D87N, gyrA-S83F, parC-S80I | 408 (7.27) |
| | gyrA-D87V, gyrA-S83F, parC-S80I | 15 (0.27) |
| | gyrA-D87G, gyrA-S83F, parC-E84K | 8 (0.14) |
| | gyrA-D87G, gyrA-S83F, parC-E84G | 1 (0.02) |
| | gyrA-D87N, gyrA-S83F, parC-E84G | 1 (0.02) |
| | gyrA-D87N, gyrA-S83F, parC-E84K | 1 (0.02) |
| | gyrA-D87G, gyrA-S83F, parC-S80I | 1 (0.02) |
| | gyrA-D87G, gyrA-S83Y, parC-S80I | 1 (0.02) |
| | gyrA-D87N, gyrA-S83F, parC-S80R | 1 (0.02) |
| **Total** | | 437 (7.79) |

**Supplementary table 4.** Estimated times of the most recent common ancestors (tMRCAs) of the main clades, with calendar years and most probable locations, from discrete phylogeography of the *S*. Typhi isolates.

| Genotype | tMCRA | Country | Substitution rate |
|---|---|---|---|
| **2.3.3** | 1817 | Vietnam | $1.55 \times 10^{-7}$ |
| **2.5** | 1856 | India | $2.85 \times 10^{-7}$ |
| **3.2.2** | 1899 | Bangladesh | $9.81 \times 10^{-8}$ |
| **3.3** | 1770 | India | $1.28 \times 10^{-7}$ |
| **4.3.1** | 1984 | India | $7.98 \times 10^{-6}$ |

### Supplementary Methods

#### Sampling of SEAP and SEFI isolates for sequencing

##### Bangladesh

A total of 820 *S*. Typhi isolates were selected for sequencing; 80 were selected for suspected resistance to azithromycin, and 740 isolates were selected from a library of 2,366 *S*. Typhi isolates using a stratified random selection procedure to sample in proportion to the distribution of samples by study year, age and sex. All isolates were collected from October, 2016 to July, 2018.

##### India

In India, isolates were obtained from 12 sites in the Surveillance for Enteric Fever in India project, which included prospective cohort studies and hospital-based surveillance. Sites were selected to be geographically representative, covering four zones (East, West, North and South). We selected 993 *S.* Typhi consecutive isolates from SEFI, obtained between October, 2017 and December, 2019.

##### Nepal

In Nepal, isolates were obtained from two studies: the Fever Etiologies Study (2014-2017) and SEAP (2017-2019). Surveillance was performed at hospitals and clinics in urban (Kathmandu), peri-urban (Kavrepalanchok) and rural areas, but the majority of typhoid cases (and isolates) were obtained from Kathmandu and Kavrepalanchok. We selected 816 randomly selected isolates spanning this study period for sequencing.

**Pakistan**

In Pakistan, isolates were obtained as part of SEAP from Aga Khan University Hospital and its affiliated laboratory network and from Kharadar General Hospital, both in Karachi. We selected a consecutive 860 isolates for sequencing, obtained between October, 2016 and December, 2018.

**Non-H58 strain selection for BEAST analyses**

Strains were selected to cover the full temporal (1905-2019) and geographical range of non-H58 lineages. For 2.3.3, 2.5, and 3.2.2 lineages, we selected all the isolates reported before our study. Isolates described in our study were selected according to the clades and branches of the maximum likelihood tree, to ensure we were including isolates with as much diversity as possible. For 3.3 lineage, we selected at least one isolate for each country and year of each sub-lineage. For countries with a large number of samples, we selected a maximum of three isolates for each country and year. From SEAP/SEFI isolates, we selected isolates according to the clades and branches of the maximum likelihood tree, with a maximum of 10 per year of each 3.3 sub-lineage.
